# Single-cell transcriptome-wide Mendelian randomization and colocalization analyses uncover cell-specific mechanisms in atherosclerotic cardiovascular disease

**DOI:** 10.1101/2024.12.19.24319316

**Authors:** Anushree Ray, Paulo Alabarse, Rainer Malik, Muralidharan Sargurupremraj, Jürgen Bernhagen, Martin Dichgans, Sebastian-Edgar Baumeister, Marios K. Georgakis

## Abstract

Genome-wide association studies (GWAS) have identified numerous genetic loci influencing human disease risk. However, linking these loci to causal genes remains challenging, limiting opportunities for drug target discovery. Transcriptome-wide association studies (TWAS) address this by linking variants to gene expression, but typically rely on bulk RNA sequencing, which lacks cell-specific resolution. Here, we present a single-cell TWAS pipeline combining *cis*-Mendelian randomization (MR) with colocalization analyses at the single-cell level. As a case study, we examined how genetically proxied gene expression in immune cells influences atherosclerotic cardiovascular disease (ASCVD) risk. We integrated single-cell expression quantitative trait loci (sc-eQTL) for 14 immune cell types with GWAS for coronary artery disease, large artery atherosclerotic stroke, and peripheral artery disease. Single-cell *cis*-MR analyses revealed 440 gene-outcome associations across cell types, 84% of which were missed by bulk TWAS, despite a considerably smaller sample size of the sc-eQTL dataset. Of these associations, 17 were replicated with external *cis*-eQTLs and demonstrated colocalization with ASCVD GWAS signals. Notably, genetically proxied expression of *LIPA* in monocytes was associated with coronary artery disease, large artery atherosclerotic stroke, and subclinical atherosclerosis traits. These findings were confirmed in a phenome-wide association study without evidence of associations with unexpected clinical outcomes. Single-cell RNA sequencing and immunohistochemistry of human carotid plaques revealed high *LIPA* expression in plaque macrophages. Our pipeline provides a solution for the discovery of cell-specific expression patterns that drive genetic predisposition to human disease, potentially impacting target selection for cell-tailored therapeutics.

## INTRODUCTION

Analyses of human genetic data can provide invaluable insights into causal disease mechanisms and inform the development of new drugs.(1) Indeed, drug targets with genetic support are more than twice as likely to deliver drugs that will be approved.(2,3) In the field of atherosclerotic cardiovascular disease (ASCVD), signals from genetic studies have informed or contributed to the emergence of several drug development programs, including PCSK9 inhibitors (4), Lp(a)-lowering molecules (5), ApoC3- and ANGPTL3-targeting agents (6), Factor XI inhibitors (7), and IL-6 signaling inhibitors.(8) Genome-wide association studies (GWAS) have identified thousands of genomic loci associated with human disease.(9) However, the translation of GWAS findings into actionable drug targets requires the determination of both causal genes regulated by the disease-associated variants and the specific cell types in which these causal genes exhibit their function.

Integrating GWAS data for clinical endpoints with data from other omics layers can provide valuable insights into causal genes for human disease at scale. For example, transcriptome-wide association studies (TWAS) use gene expression levels instrumented by *cis*-expression quantitative trait loci (*cis*- eQTL) to identify tissue-specific and functionally relevant genes associated with disease outcomes from GWAS loci.(10) However, gene expression is regulated at the cellular level and not the tissue level. As such, eQTLs could be specific to distinct cell types that are relatively rare in a given tissue and obscured in bulk analyses that average gene expression from diverse cell types. A higher-resolution characterization of the biological complexity and cellular heterogeneity of ASCVD could be obtained from single-cell omics technologies. Integration of single-cell-transcriptome profiles from single-cell RNA sequencing (scRNA-seq) and GWAS data could enhance our current understanding of disease mechanisms, aid the identification of cell-specific druggable targets, and facilitate the development of tailored interventions, such as cell-targeted RNA therapeutics.

Here, we present a single-cell TWAS pipeline that combines *cis*-Mendelian randomization (MR) using cell-specific *cis*-eQTL variants alongside colocalization analyses to identify causal cell-specific expression changes underlying GWAS signals. As a case study, we integrated immune cell-specific eQTL data with GWAS summary statistics for the most common manifestations of ASCVD — coronary artery disease (CAD), large artery atherosclerotic stroke (LAS), peripheral artery disease (PAD) — to explore potential cell-specific immune mechanisms involved in atherosclerosis. Our analysis revealed both established and novel causal genes that could not be captured using bulk TWAS analyses, uncovering both the specific cell types in which these genes might exhibit their effects, as well as the direction of their effects on ASCVD outcomes. Using cell-specific *cis*-eQTL data from external cohorts, we replicated significant findings. We further performed downstream experimental and computational analyses to investigate an association between higher genetically proxied *LIPA* expression in monocytes and atherosclerosis.

## RESULTS

### Overview of the method and data sources

Our proposed pipeline for a single-cell TWAS study is summarized in **Figure 1**. Briefly, similar to previous TWAS approaches at bulk level (11–13), we use single-cell *cis*-eQTLs derived from scRNA-seq studies as instruments for downstream MR analyses in GWAS summary data for outcomes of interest (discovery MR). As opposed to bulk RNA-seq, scRNA-seq datasets are usually smaller in scale. Therefore, to minimize false positive rates, we use an external dataset for the selection of single-cell *cis*- eQTLs for replication of the MR results (replication MR). Finally, to minimize confounding due to the pleiotropic effects of the variants on the expression of neighboring genes, we use colocalization analyses.

**Figure 1.**
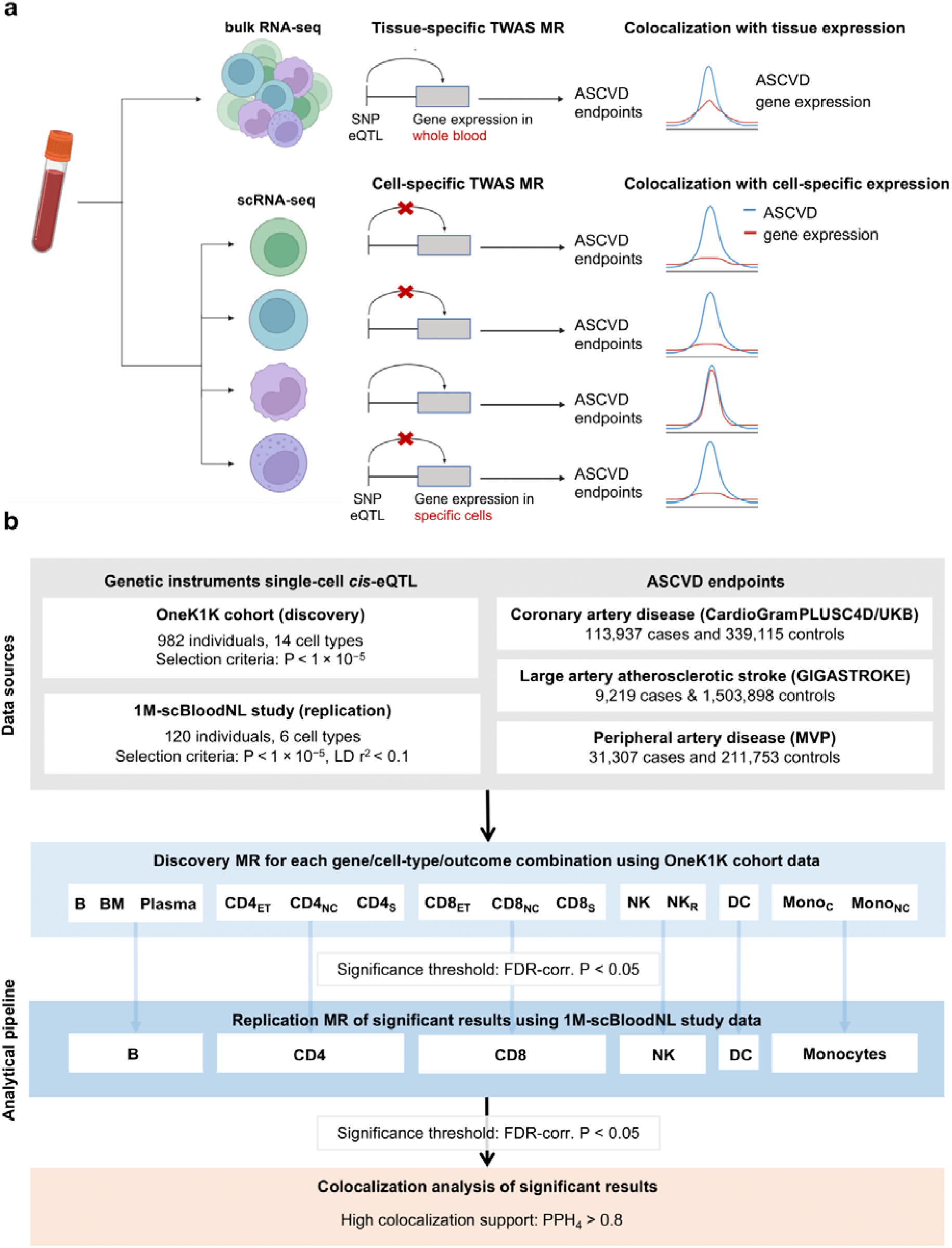
Study design. (a) Schematic of our proposed approach for transcriptome-wide association studies using single- cell RNA sequencing (RNA-seq) vs. bulk RNA-seq data. Our approach includes Mendelian randomization analyses followed by colocalization. (b) Overview of the integrative genomic analysis pipeline and data sources used for this study. eQTL, expression quantitative trait loci; GWAS, genome-wide association study; CAD, coronary artery disease; LAS, large artery stroke; PAD, peripheral artery disease; MR, Mendelian randomization; B, Immature and naïve B cell; BM, Memory B cell; CD4NC, CD4+ naïve and central memory T cell; CD4ET, CD4+ effector memory and central memory T cell; CD4S, CD4+ SOX4 T cell; CD8NC, CD8+ naïve and central memory T cell; CD8ET, CD8+ effector memory T cell; CD8S, CD8+ S100B T cell; NK, Natural killer cell; NKR, Natural killer cell Recruiting; MonoC, Classical monocyte; MonoNC, Non-classical monocyte; DC, Dendritic cell.; FDR, false-discovery rate; PPH, posterior probability hypothesis; TWAS, transcriptome-wide association study.

In our case study, we focused on exploring immune cell-specific mechanisms driving the risk of ASCVD, given the accumulating interest in the role of immune mechanisms in atherosclerosis.(14) We started by leveraging cell-specific *cis*-eQTL data from peripheral blood mononuclear cells (PBMCs) in the OneK1K cohort (N=982). In MR analyses, we then explored the effects of genetically proxied immune cell-specific gene expression on CAD (113,937 cases and 339,115 controls), LAS (9,219 cases and 1,503,898 controls), and PAD (31,307 cases and 211,753 controls) using the largest available publicly available GWAS summary datasets. We replicated significant findings in single-cell *cis*-eQTLs data from the 1M-scBloodNL study (N=120) and then applied colocalization analyses between *cis*-eQTL data from the OneK1K study and GWAS statistics for ASCVD phenotypes.

### Single-cell TWAS-MR uncovers cell-specific gene expression effects on ASCVD not captured by bulk TWAS-MR

Of the 6,468 genes analyzed in the OneK1K cohort, 5,162 (79.8%) had significant *cis*-eQTLs. The number of genes with significant *cis*-eQTLs varied widely across cell types, ranging from 4,411 for CD4+ naive and central memory T cells (CD4_NC_) cells to 244 for plasma cells. For the majority of gene/cell-type combinations (81.7%), only a single *cis*-eQTL was retained as instrument (**Supplementary Figure 1** and **Supplementary Tables 1 – 3)**. Between 7% to 61% of *cis*-eQTLs detected in individual cell types were not detected as *cis*-eQTL in bulk RNA sequencing of whole blood in the much larger dataset of the eQTLGen consortium (N=31,686 for bulk *cis*-eQTL vs. N=982 for single-cell *cis*-eQTL; **Supplementary Figure 2A, B**).

Detailed results of the discovery MR analysis (Wald ratio MR when the instrument consisted of a single *cis*-eQTL or inverse-variance weighted MR when the instrument consisted of >1 *cis*-eQTLs) examining the relationship between genetically proxied cell-specific gene expression and ASCVD outcomes are shown in **Supplementary Tables 4 – 6**. Of 34,347 gene/cell-type/outcome combinations analyzed, 440 showed significant MR effect estimates (false discovery rate [FDR]-corrected p-value < 0.05), representing 318 unique gene-outcome pairs across different cell types. Notably, only 52 (16.4%; 32% for CAD, 3.9% for LAS, 11.2% for PAD) of these significant associations were captured in MR analyses using bulk *cis*-eQTLs for whole blood, despite the much larger sample size used for *cis*-eQTL detection in bulk RNA sequencing (N=31,686 vs. N-982; **Figure 2**). Furthermore, across all MR results, there were low to moderate correlations between bulk and single-cell effect estimates for all outcomes (median Pearson’s *r* for CAD 0.49 [range 0.02 – 0.75], for LAS 0.43 [range -0.11 – 0.7], and for PAD 0.34 [range- 0.33 – 0.7]) (**Figure 2**). Collectively, these results indicate a significant gain in identified signals from the single-cell TWAS MR approach compared to bulk TWAS MR despite the considerably smaller sample sizes for single-cell *cis*-eQTL discovery datasets.

**Figure 2.**
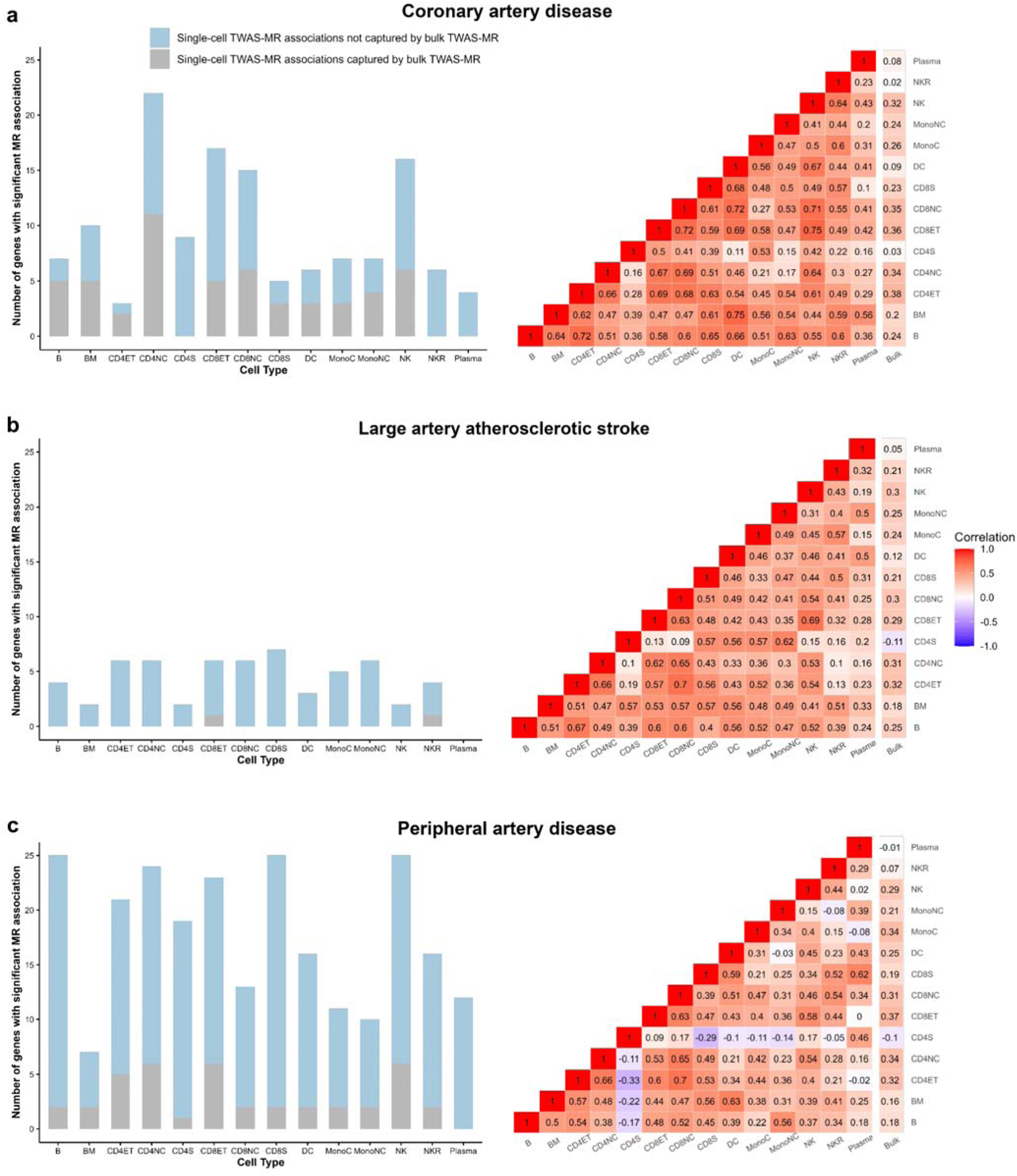
Comparison between single-cell and bulk Mendelian randomization analyses results. Stacked bar plot and correlation matrix of Mendelian randomization estimates for different immune cell types vs. whole blood for (a) coronary artery disease, (b) large artery atherosclerotic stroke, and (c) peripheral artery disease B, Immature and naïve B cell; BM, Memory B cell; CD4NC, CD4+ naïve and central memory T cell; CD4ET, CD4+ effector memory and central memory T cell; CD4S, CD4+ SOX4 T cell; CD8NC, CD8+ naïve and central memory T cell; CD8ET, CD8+ effector memory T cell; CD8S, CD8+ S100B T cell; NK, Natural killer cell; NKR, Natural killer cell Recruiting; MonoC, Classical monocyte; MonoNC, Non-classical monocyte; DC, Dendritic cell.

### Replication of single-cell TWAS-MR and colocalization analyses

Of the 440 significant gene/cell-type/outcome combinations identified in the discovery MR, 38 achieved an FDR-corrected p-value < 0.05 in the replication MR analysis using genetic instruments from the 1M- scBloodNL study (**Figure 3A** and **Supplementary Tables 7, 8**). Of these, 17 gene/cell type/outcome combinations (14 unique genes) were also supported by a high posterior probability for a shared causal genetic variant in colocalization analyses (PPH4[≥[0.8, **Figure 3A** and **Supplementary Table 9**). The effects of the 14 genes on the three outcomes across different cell types are shown in **Figure 3B**.

**Figure 3.**
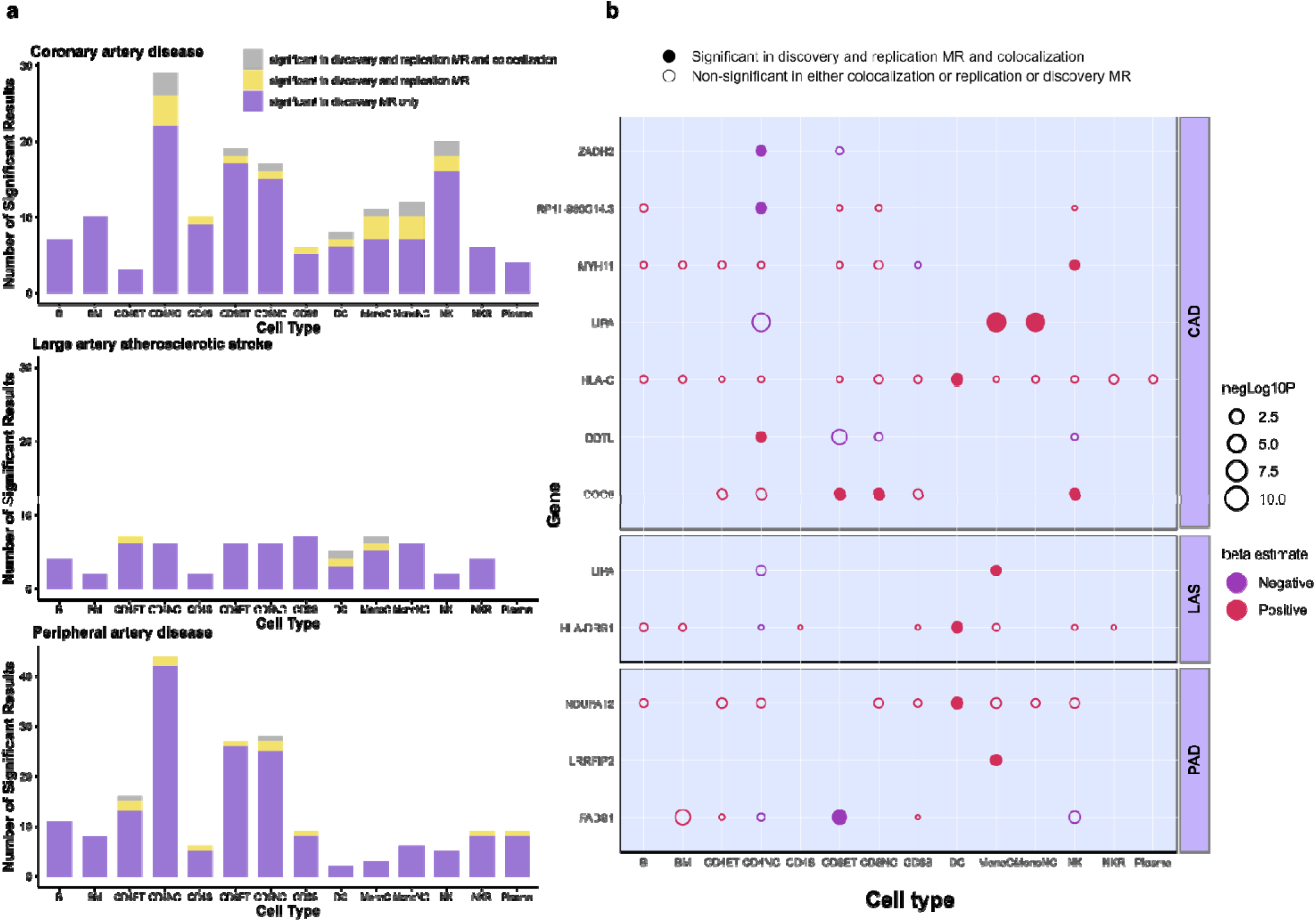
Associations between immune cell-type-specific gene expression and atherosclerotic cardiovascular disease outcomes. (a) Stacked bar graphs of the number of genes whose cell-specific expressions were found to be significantly associated with coronary artery disease, large artery atherosclerotic stroke, and peripheral artery disease in each step of the statistical analysis. (b) Bubble heatmaps for cross-cell-type comparison of discovery Mendelian randomization (MR) estimates of cell-type/gene/outcome combinations which had robust MR and colocalization evidence. Filled bubbles indicate associations that were significant in each of the three steps of the analysis pipeline. The color of the bubble corresponds to the beta coefficient of the association between the genetically predicted expression of genes (y-axis) across different cell types (x-axis) and the disease outcome. The size of each bubble corresponds to the negative logarithm of the discovery MR association false discovery rate-corrected p-value. B, Immature and naïve B cell; BM, Memory B cell; CD4NC, CD4+ naïve and central memory T cell; CD4ET, CD4+ effector memory memory T cell; CD4S, CD4+ SOX4 T cell; CD8NC, CD8+ naïve and central memory T cell; CD8ET, CD8+ effector memory T cell; Cnd central 8S, CD8+ S100B T cell; NK, Natural killer cell; NKR, Natural killer cell Recruiting; MonoC, Classical monocyte; MonoNC, Non-classical monocyte; DC, Dendritic cell; ZADH2, zinc-binding alcohol dehydrogenase domain-containing protein 2; RP11-950C14.3, lncRNA, antisense to EIF2B ; MYH11, myosin heavy chain 11; LIPA, Lipase A, lysosomal acid type; HLA, human leukocyte antigen; DDTL, D-dopachrome tautomerase like; COG5, component of oligomeric golgi complex 5; NDUFA12, NADH:ubiquinone oxidoreductase subunit A12; LRRFIP2, leucine-rich repeat flightless- interacting protein 2; FADS1, fatty acid desaturase 1; LAS, large artery stroke; CAD, coronary artery disease; PAD, peripheral artery disea e.

Although the genetically proxied expression of several genes, including *ZADH2*, *HLA-C*, *COG5*, *NDUFA12,* and *LRRFIP2,* had directionally consistent effects, many genes had cell-specific effects. Specifically, higher *LIPA* expression in monocytes was associated with an increased risk of CAD and LAS, whereas higher *LIPA* expression in CD4_NC_ cells was associated with a lower risk of CAD and LAS. Higher *DDTL* expression was consistently associated with a lower risk of CAD when expressed in CD8+ effector memory T cells (CD8_ET_), CD8+ naïve and central memory T cells (CD8_NC_), and Natural killer (NK) cells, but its expression in CD4_NC_ cells was associated with a higher risk of CAD. Additionally, higher expression of *MYH11* and *RP11-950C14.3* was associated with lower risks of CAD only in CD8+S100B T cells (CD8_S_) and CD4_NC_ cells, respectively. Finally, higher expression of *FADS1* was associated with increased risk of PAD in Memory B cells (BM), CD4+ effector memory and central memory T cells (CD4_ET_), and CD8_S_ cells, and decreased risk of PAD in CD4_NC_, CD8_ET_, and NK cells.

### Genetically proxied monocyte expression of *LIPA* associated with atherosclerosis

The colocalization between *LIPA* eQTLs and CAD and LAS GWAS association signals was specific for monocytes (**Figure 4A**). To further investigate this signal and explore associations with outcomes other than ASCVD, we performed a phenome-wide association study (PheWAS) analysis on 487,314 participants of the UK Biobank. After correcting for multiple comparisons (FDR-corrected p-value < 0.05), the only phenotypes significantly associated with higher genetically proxied *LIPA* expression in the PheWAS analyses were myocardial infarction, coronary atherosclerosis, and ischemic heart disease, thereby validating the relevance of *LIPA* in monocytes for ASCVD in an external dataset (**Figure 4B** and **Supplementary Table 10)**. There was no evidence of associations with other phenotypes in the opposite direction, supporting a favorable safety signal when genetically perturbing this drug target. Beyond clinical endpoints, we also found a significant association between genetically proxied monocyte *LIPA* expression and carotid plaque as captured by ultrasound, as well as myocardial infarction, ischemic heart disease, and coronary atherosclerosis (**Figure 4C**).

**Figure 4.**
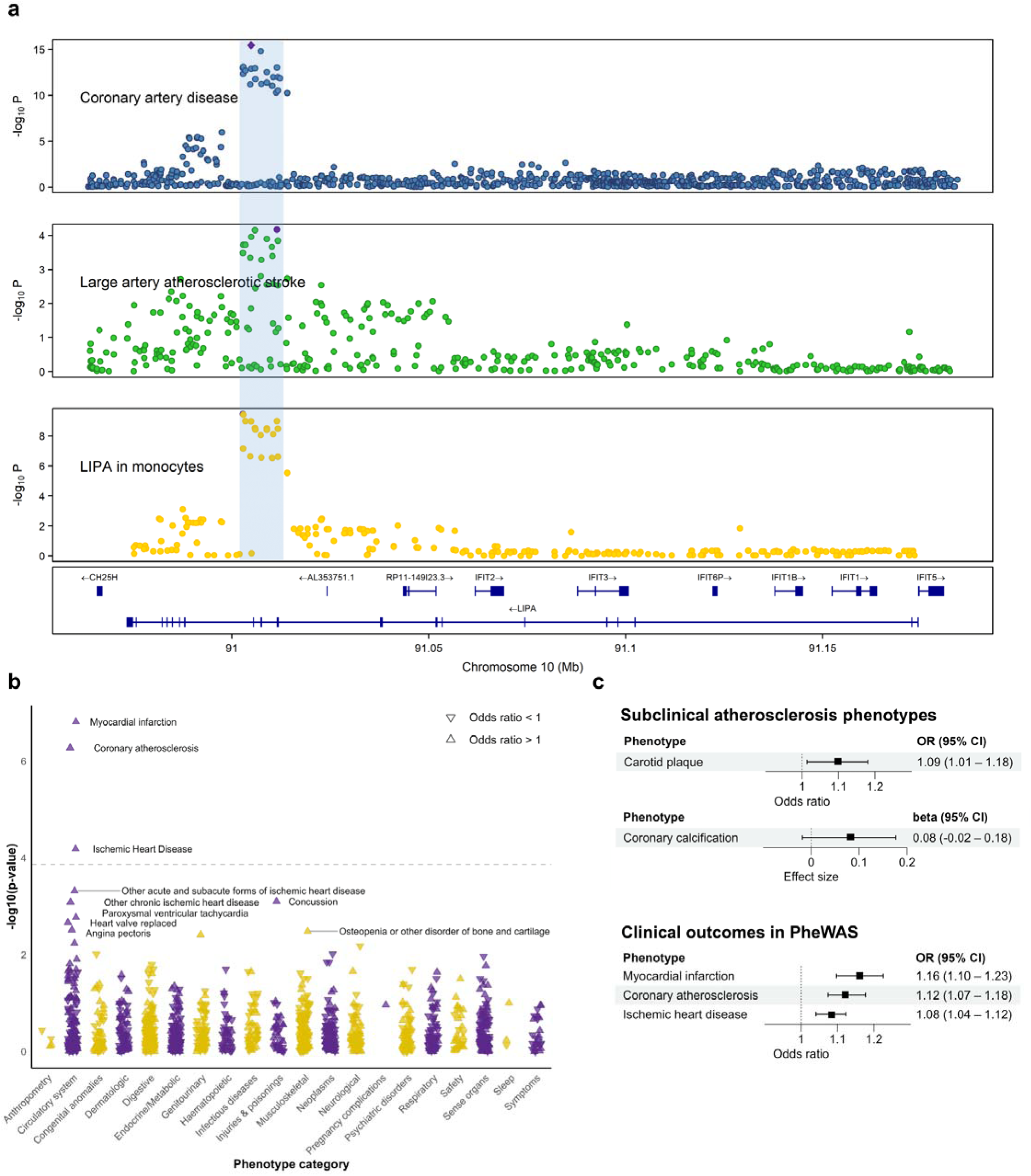
Association between genetically proxied monocyte expression of *LIPA* and atherosclerotic cardiovascular disease outcomes. (a) LocusZoom plots illustrating evidence of genetic colocalization between *LIPA* gene expression in monocytes and coronary artery disease and large artery atherosclerotic stroke in the *LIPA* gene locus. (b) Forest plots of Mendelian randomization (MR) results for the effects of genetically proxied *LIPA* expression in monocytes and atherosclerotic cardiovascular outcomes. (c) Manhattan plot of a MR-phenome-wide association study for genetically proxied *LIPA* expression in monocytes. The dashed horizontal gray line represents the false discovery rate-corrected p- value of 0.05.

### mRNA expression and protein levels of *LIPA* in human atherosclerotic plaque macrophages

Given the evidence for an effect of monocyte-specific expression of *LIPA* on ASCVD, we, in a last step, examined whether *LIPA* is expressed in human atherosclerotic plaques and, more specifically, in plaque macrophages, which are primarily derived from circulating monocytes. Using published scRNA-seq data from 15 advanced human carotid artery plaques (**Supplementary Figure 3**),(15) we found *LIPA* to be expressed throughout all detected cell types, but its expression was highest in macrophages (**Figure 5A**). Accordingly, immunohistochemical staining of human carotid plaques from 3 patients undergoing endarterectomy from the AtherOMICS cohort demonstrated LIPA in CD68-stained macrophages along with abundant cholesterol clefts (**Figure 5B** and **Supplementary Figure 4**).

**Figure 5.**
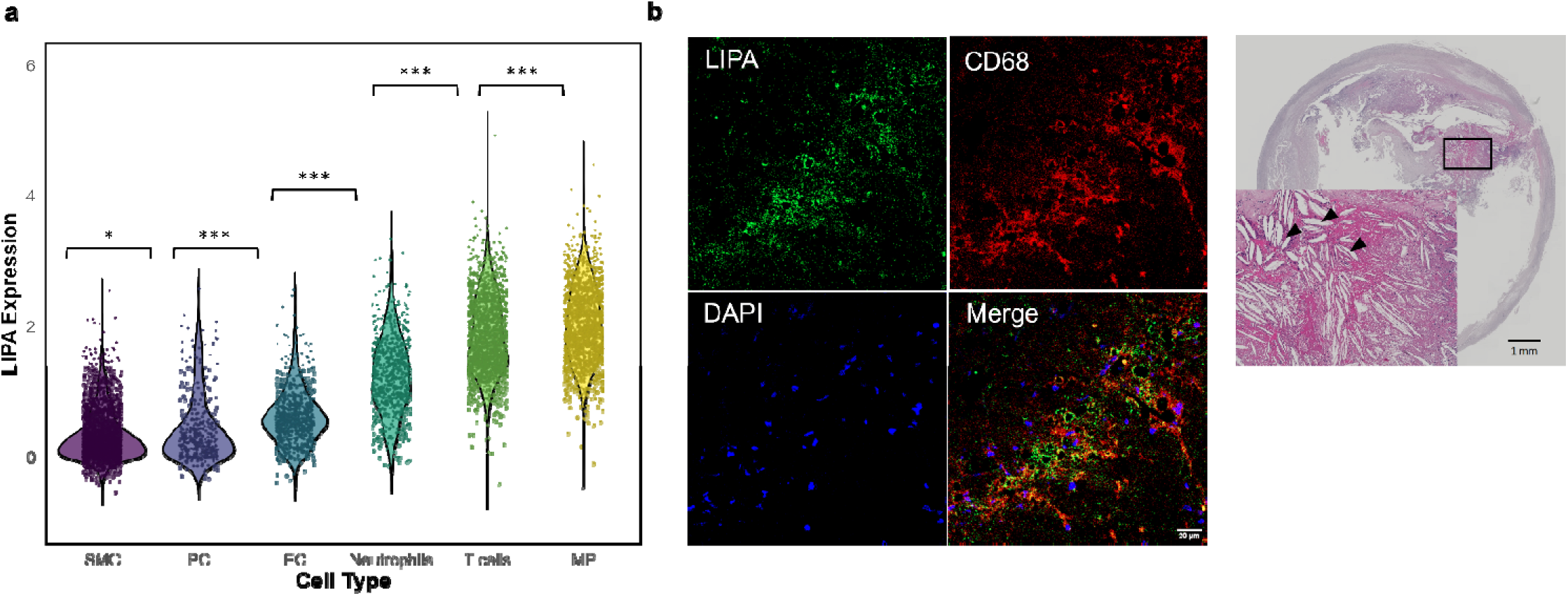
*LIPA* expression in human carotid atherosclerotic plaques. (a) Violin plot of *LIPA* expression across different cell types in 15 human atherosclerotic plaque samples from Mocci et al.(15) P-values of Wilcoxon rank sum test of expression between cell types are indicated as ***, p-value < 0.001; **, p-value < 0.01; *, p-value < 0.05. (b) Cholesterol clefts (black arrowheads) and LIPA in macrophages (CD68) in a human carotid plaque section from a symptomatic patient (n =3) SMC, smooth muscle cells; PC, pericytes; EC, endothelial cells; MP, macrophages.

## DISCUSSION

In the present study, we proposed and implemented an analytical pipeline for TWAS at the single-cell level. We demonstrated the applicability and potential of the approach by integrating single-cell *cis*-eQTL data for 14 immune cell types in peripheral blood with GWAS data for three ASCVD outcomes — CAD, LAS, PAD. Our single-cell MR analyses revealed significant information gains compared to bulk TWAS MR, despite considerably smaller sample sizes of the single-cell *cis*-eQTL discovery datasets. A total of 17 associations between cell-specific gene expression and ASCVD outcomes passed our stringent screening criteria, which included replication using sc-eQTLs derived from an alternative scRNA-seq dataset and genetic colocalization evidence between cell-specific expression and ASCVD outcomes. While the expression of genes such as *MYH11* and *COG5* was found to affect disease risk in multiple immune cell types, higher genetically proxied expression of *LIPA* was associated with a higher risk of two ASCVD outcomes — CAD and LAS — specifically in monocytes. We validated the associations of monocytic expression of *LIPA* with atherosclerotic endophenotypes and clinical endpoints in PheWAS analyses in an external dataset, which also broadly supported a favorable safety profile with no significant signals for higher risk of unexpected clinical outcomes. Finally, follow-up analyses of scRNA-seq data from human carotid plaques revealed high expression of *LIPA* in plaque macrophages, which was also confirmed at the protein level through immunohistochemistry.

Our approach enhances the conventional bulk TWAS-MR paradigm, enabling the detection of cell-specific expression patterns driving genetic predisposition to human disease. Since most genetic polymorphisms associated with human diseases are located in non-coding regions, it is believed that genetic variation influences predisposition to disease primarily by influencing gene expression patterns.(16) In this context, TWAS explorations integrating GWAS findings with tissue-specific eQTL data have become crucial in post-GWAS explorations.(17–19) As gene expression is regulated at the cellular level, using sc-eQTL data has significant benefits. Although scRNA-seq studies are still limited by sample size, we found that using cell-specific eQTL instruments enables the detection of signals that would not be detected with bulk eQTL instruments derived from much larger studies. This is reflected in the relatively weak correlation between cell-specific MR and bulk MR estimates for all coded genes across ASCVD outcomes. By integrating scRNA-seq eQTL data with GWAS risk loci using MR and colocalization analyses, we obtained evidence of potentially causal genes at risk loci for ASCVD and resolved specific cell types through which these genes exert their pathogenetic effects. For example, increased CD4_NC_ cell-specific *DDTL* expression was found to be associated with higher CAD risk, whereas increased Mono_C_-specific *LRRFIP2* expression was found to be associated with higher PAD risk. Our approach is generalizable to other outcomes, as well as single-cell and single-nuclei RNA-seq data from tissues beyond peripheral blood, as such data become increasingly available.

Higher genetically proxied monocyte expression of *LIPA* was associated with a higher risk of both CAD and LAS, highlighting its role in the development of atherosclerosis. The *LIPA* locus has been consistently identified as a risk locus for CAD in previous GWAS, with colocalization analyses suggesting *LIPA* as the causal gene at this locus.(20,21) CAD risk-enhancing alleles are also eQTLs for *LIPA* expression in whole blood.(22) Our findings provide robust evidence that the effect of these genetic variants on *LIPA* expression in monocytes, rather than other PBMCs, drives the risk of CAD, LAS and other atherosclerotic phenotypes. This aligns with a previous *in vitro* study in isolated human monocytes, which showed that risk-enhancing alleles not only increase *LIPA* expression but also the activity of the LIPA enzyme.(23) In line with this, a study in *Ldlr* knockout mice has demonstrated that myeloid-specific *Lipa* overexpression leads to larger atherosclerotic lesions with higher macrophage content.(24) *LIPA* encodes lysosomal acid lipase, a key enzyme in lipid metabolism that hydrolyzes cholesteryl esters and triglycerides in lysosomes.(25) Given that most atherosclerotic plaque macrophages originate from circulating monocytes, it is plausible that LIPA exerts its risk-enhancing effect by promoting excessive cholesterol crystal formation within these lesion macrophages. Supporting this hypothesis, our analysis revealed high LIPA mRNA and protein levels in macrophages within human atherosclerotic plaque samples with abundant cholesterol crystals. On the other hand, rare loss-of-function mutations leading to complete loss of LIPA activity and partial residual activity cause infant-onset Wolman disease and cholesteryl ester storage disease, respectively, with the latter also being associated with premature atherosclerosis, likely due to severe hyperlipidemia.(26,27) This dual role—where reduced LIPA activity leads to hyperlipidemia-driven atherosclerosis and elevated LIPA expression may drive pro-inflammatory actions in macrophages—highlights the importance of LIPA homeostasis in preventing atherosclerosis. Reciprocally, the ability of our analysis pipeline to identify LIPA as an ASCVD risk driver, further supported by our PheWAS-MR and atherosclerotic plaque RNA sequencing analysis, reinforces the robustness of our approach in detecting cell-specific genetic drivers of ASCVD risk.

The findings from our study could have implications for the development of RNA-based therapeutics, particularly those targeting gene expression in specific cell types. The identification of cell-specific gene expression patterns, such as the association of *LIPA* expression in monocytes and macrophages with ASCVD, underscores the potential for designing cell-tailored RNA therapies. RNA-based drugs are gaining pace in the cardiovascular field and becoming increasingly common, demonstrating the feasibility and effectiveness of these modalities. RNA-based drugs have been successfully introduced to clinical practice, such as inclisiran, a silencing RNA (siRNA) agent that targets the synthesis of PCSK9 to lower LDL cholesterol (28), or are in advanced stages of clinical development, such as siRNA therapeutics against APOC3, ANGPTL3, Lp(a), and angiotensinogen.(29–32) By focusing on modulating gene expression within specific cell types, it may be possible to mitigate disease risk while minimizing off-target effects. As RNA-based drugs continue to advance, our sc-TWAS MR pipeline provides a robust framework for *in silico* identification and validation of cell-specific targets, paving the way for more effective and safer therapies for complex diseases like ASCVD.

Our study has limitations. First, MR analysis at the single-cell level may miss risk genes with lower expression levels because the sparse expression in individual cells can limit statistical power, whereas bulk analysis averages gene expression across many cells, improving the detection of lowly expressed genes. Second, the relatively small sample size of scRNA-seq studies limited the number of genes tested and the number of eQTLs detected. The number of eQTLs obtained for each cell type from publicly available datasets varied according to the sample sizes for eQTL analyses of each cell type. Since most genes were associated with only one eQTL as valid IV, sensitivity analyses to correct for horizontal pleiotropy could not be performed. The follow-up colocalization analyses provided, however, evidence for a shared genetic basis between the exposure and outcome rather than horizontal pleiotropy. Third, the default coloc assumption of a single causal variant per locus could have led to an underestimation of colocalization in loci with multiple causal variants. Fourth, differences in scRNA-seq protocols could cause substantial variability in results. However, the consistency of the main findings across two independently collected scRNA-seq datasets raises confidence in their validity, although the finer resolution of cell types in the discovery dataset compared to the replication dataset may limit their direct comparability. Fifth, the GWAS and eQTL analyses in this study were primarily conducted on individuals of European ancestry, which may limit the generalizability of the findings to other ethnicities. Sixth, due to the lack of sc-eQTL data from human vasculature, we could not apply our transcriptome-wide approach to potentially more relevant tissues, where many of the ASCVD-associated variants might exert their effects.

In conclusion, we propose an integrative single-cell TWAS pipeline that could enhance our understanding of cell-specific gene expression patterns driving genetic predisposition to human disease. Using results from this approach as a foundation, we provided support for the key role of monocyte-specific *LIPA* in atherosclerosis with potential therapeutic relevance. Findings from single-cell TWAS could inform target selection for therapeutic modalities tailored to specific cell types, such as RNA therapeutics.

## METHODS

### Study design

The study design is summarized in **Figure 1**. Briefly, we used whole blood sc-eQTL data from circulating immune cells and GWAS summary statistics for different ASCVD outcomes to conduct discovery and replication MR analyses. Thereafter, we analyzed significant target genes for each cell type and ASCVD outcome with colocalization analysis.

### Selection of genetic instruments

We obtained sc-eQTL mapping data, which integrate genotyping and scRNA sequencing data, from the OneK1K cohort for the discovery analyses and the 1M-scBloodNL study for the replication analyses.(33,34) The OneK1K cohort generated data from 1,267,758 PBMCs from 982 healthy individuals of Northern European ancestry.(33) Genotyping data were imputed using the Michigan Imputation Server with the Haplotype Reference Consortium panel and scRNA-seq data were generated using a pooled multiplexing strategy. Through unsupervised clustering of their transcriptional profile with Seurat (35), 14 cell types were defined: B cell lineage was classified as plasma cells, immature and naive B cells, or memory B cells. CD4+ T cells were classified as naive and central memory cells (CD4_NC_), effector memory and central memory T cells (CD4_ET_), and SOX4-expressing T cells (CD4_SOX4_). Similarly, CD8+ T cells were classified as CD8_NC_, CD8_ET_, and CD8_SOX4_ cells. Innate immune lymphocytes were distinguished into NK and NK recruiting cells, classical and non-classical monocytes, and dendritic cells. For each gene/cell type combination, *cis*-eQTLs were identified using Spearman’s rank correlation testing between the gene expression levels and SNPs within a 1000-kb region of either end of the gene. The top five independent eQTLs were identified by selecting the SNP with the smallest FDR-corrected p-value (eSNP_1_) in the correlation test and then performing five rounds of iterative conditional analysis to yield eSNP_1_ to eSNP_5_. Summary statistics were provided for all five SNPs for each gene/cell type combination. For each gene/cell-type combination, we only used eQTLs reaching a P-value threshold of <[1[×[10^−5^, corresponding to an FDR-corrected P < 0.05 in a previous TWAS analysis.(10) The 1M-scBloodNL study generated data from 928,275 PBMCs from 120 individuals from the Northern Netherlands population cohort Lifelines. Genotyping data were imputed using the Michigan Imputation Server with the Haplotype Reference Consortium panel and scRNA-seq data were generated using a pooled multiplexing strategy. Sc-eQTL mapping data were available for six cell types based on marker gene expression: B cells, CD4+ T cells, CD8+ T cells, monocytes, NK cells, and DC cells. For each gene/cell type combination, summary statistics for associations of all SNPs within a 100 kb distance from the gene midpoint encoding the respective transcript were generated and were available for our analyses. We selected *cis*-eQTLs on the basis of an association at a P[<[1[×[10^−5^ and clumped them for linkage disequilibrium (LD) using the clump_data function at a threshold of r^2^ < 0.1.

To compare our approach to a conventional TWAS, bulk eQTL summary statistics from 31,684 whole blood samples of mostly European ancestry were obtained from the eQTLGen Consortium.(36) We selected as genetic instruments *cis*-eQTLs within a 100 kb distance from the gene midpoint encoding the respective transcript that was associated with the levels of the respective transcript at a P[<[1[×[10^−5^. Thereafter, we clumped the genetic variants for LD at a threshold of r^2^ < 0.1.

### Clinical endpoints and intermediate phenotypes

We obtained GWAS summary statistics for ischemic stroke and its subtypes from the GIGASTROKE trans-ancestry GWAS meta-analysis of 86,668 cases (67% European, East Asian, African, Hispanic, and South Asian ancestries) and 1,503,898 controls.(37) Of the ischemic stroke cases, 9,219 were subclassified as LAS. Summary statistics for CAD were obtained from a GWAS meta-analysis of 113,937 cases and 339,115 controls of mostly (>95%) European ancestry conducted by Nelson et al.(38) Summary statistics for PAD were obtained from a GWAS conducted in the Million Veteran Program (31,307 cases and 211,753 controls) of European, African, and Hispanic ancestry.(39) For follow-up analyses, we also used data for atherosclerosis endophenotypes –carotid plaque and coronary calcification. We obtained summary statistics from GWAS meta-analyses of cohorts of the CHARGE Consortium, including 48,434 individuals of European ancestry for carotid plaque (21,540 cases, 26,894 controls) and 35,776 individuals of primarily (75%) European ancestry for coronary artery calcification score.(40,41)

### Mendelian randomization

We undertook a two-stage (discovery and replication) MR approach to systematically evaluate evidence for the putative causal effects of immune cell-specific gene expression on the six cardiovascular outcomes.(10) The discovery MR analyses were conducted between *cis*-eQTLs in 14 immune cell types from the OneK1K cohort and CAD, LAS, and PAD using the TwoSampleMR R package (v 0.6.8).(42) To ensure the identical orientation of effect alleles between the eQTL and outcome associations, we harmonized the exposure and outcome datasets using the harmonise_data() function. Subsequently, if only a single eQTL was available for a gene, the Wald ratio estimate was obtained. If more than one SNP was available, the inverse-variance weighted (IVW) method was used to obtain an effect estimate. All P-values were adjusted using the Benjamini-Hochberg method for controlling the False Discovery Rate (FDR) in multiple comparisons.(43) For each outcome, pairwise weighted Pearson correlation of the discovery MR analyses results between different cell types, as well as bulk-eQTL MR analyses results, were determined and visualized as a correlation matrix.

We brought forward significant target genes in the discovery MR analyses (FDR-corrected p[<[0.05) to the replication MR analysis. We reclassified the 14 cell types from the OneK1K cohort to the less dimensional six cell types in the 1M-scBloodNL study — B cell, CD4+ T cell, CD8+ T cell, NK cell, monocyte, and dendritic cell.

### Colocalization

For significant cell-type/gene/outcome MR associations (FDR-corrected p-value[<[0.05) in the replication MR analyses, we additionally performed colocalization analysis to determine whether gene expression and cardiovascular outcomes shared the same causal variant rather than the variant being shared due to LD. Colocalization analysis provides the posterior probabilities (PP) of five hypotheses: neither gene expression nor the outcome is associated with genetic variants in the region (H0), only gene expression is associated with a genetic variant in the region (H1), only the outcome is associated with a genetic variant in the region (H2), gene expression and outcome are both associated with the region, but with different causal variants (H3), gene expression and outcome are associated with the same causal variant (H4). We performed colocalization analysis using the Bayesian coloc.abf function in the coloc R package (v 5.2.3) with default prior probabilities p1 = p2 = 1 × 10^−4^, p12 = 1 × 10^-5^.(44) Significant colocalization was defined as PPH4[≥[0.8.(44)

### Phenome-wide association study

To test the association between genetically proxied *LIPA* expression in monocytes with the full range of clinical phenotypes and detect possible unexpected associations with unexplored phenotypes, we used DeepPheWAS and assigned 487,314 participants from the population-based UK Biobank (UKB) to standardized Phecodes representing disease entities.(45) We used all ICD10 codes (main position, secondary position, death records) from the UKB. We excluded Phecodes with <100 cases and Phecodes that are male- or female-specific, leading to a total of 1,312 phenotypes. Individuals were assigned a case status if >1 ICD10 code mapped to the respective Phecode. Individuals meeting the pre-specified exclusion criteria were removed from the analysis; otherwise, the individual was assigned a control status. We used logistic regression with age, sex, and 10 principal components as covariates to test variant carrier status (0/1) against the phenotype of interest. Wald ratio MR analyses were performed for genetically proxied monocyte *LIPA* expression (1 variant). Results reaching an FDR-corrected p-value < 0.05 were considered statistically significant.

### Single-cell RNA sequencing analysis in human atherosclerotic plaques

To explore the expression of *LIPA* beyond whole blood in human atherosclerotic lesions, we downloaded individual-level scRNA-seq data from 15 carotid atherosclerotic plaques from Mocci et al.(15) We analyzed the raw count matrices using the Seurat pipeline (v 5.1.0).(35) In the initial preprocessing, we filtered out cells with fewer than 300 detected genes, those with total gene counts outside the range of 50,000 to 750,000, and cells with mitochondrial gene content exceeding 10% of total gene expression. Thereafter, we performed data normalization, identification of variable features, and scaling. To integrate data across samples, we selected common features and applied principal component analysis (PCA) to each dataset. We combined datasets using integration anchors, followed by additional PCA and uniform manifold approximation and projection (UMAP) for dimensionality reduction and clustering. We annotated clusters by comparing cluster-specific marker genes with known cell-type markers and renamed cluster identities to accurately reflect cell types. A UMAP plot was generated to visualize the integrated data.

### Immunohistochemistry for LIPA in human atherosclerosis plaques

Carotid plaque samples were obtained from patients undergoing carotid endarterectomy at the Department of Vascular Surgery of the LMU University Hospital in Munich. The AtherOMICS biobank has been approved by the Ethics Commission at the LMU Munich (approval no. 22-0135) and was conducted according to the Declaration of Helsinki. Written informed consent was obtained from each patient. Following removal of the plaque, the carotid samples were fixed in 4% paraformaldehyde+0.1 M PBS (pH 7.4) for 24 h, decalcified in EDTA (200 mM EDTA, 50 mM Trizma Base, pH 8.0), dehydrated, embedded in paraffin, and sectioned into 3.5 μm sections with a microtome. Plaque sections from three symptomatic patients were used for staining. Slides were deparaffinized with Roti-Histol, then rehydrated progressively from 100% ethanol to distilled water, 5 min incubation in each step. Then, after antigen retrieval with trypsin for 10 min at 37°C, permeabilization in Tris-buffered saline (20 mM Trizma base, 200 mM sodium chloride, pH 7.6) + 0.025% Triton X-100, pure cold methanol fixation for 10 min, and blocking with 2.5% normal horse Serum for 20 min, sections were incubated with primary antibodies against LIPA (1:50; PA5-97928, ThermoFisher) and CD68 (1:100; 14-0681-82, ThermoFisher) overnight at 4°C and fluorescent secondary antibodies (VectaFluor Duet Immunofluorescence Double Labeling Kit, DyLight™ 488 Anti-Rabbit, DyLight™ 594 Anti-Mouse, Vectorlabs) for 1 h at room temperature. Sections were mounted with DAPI (Abcam) mounting media with antifade agent (Vectashield, Vectorlabs). Image acquisition was performed using a confocal microscope (LSM 980, Carl Zeiss), and images were recorded and processed with ZEN software (Carl Zeiss, version 3.3).

### Data availability

All data produced in the present work are contained in the manuscript We performed analyses using publicly available data obtained from links provided below: Sc-eQTL summary statistics for discovery analysis: https://www.science.org/doi/10.1126/science.abf3041 Sc-eQTL summary statistics for replication analysis: https://eqtlgen.org/sc/datasets/1m-scbloodnl-eqtls.html Bulk eQTL summary statistics: https://www.eqtlgen.org/cis-eqtls.html Cross-ancestry summary statistics for LAS: http://ftp.ebi.ac.uk/pub/databases/gwas/summary_statistics/GCST90104001-GCST90105000/GCST90104538/ Summary statistics for CAD: https://www.cardiogramplusc4d.org/media/cardiogramplusc4d-consortium/data-downloads/UKBB.GWAS1KG.EXOME.CAD.SOFT.META.PublicRelease.300517.txt.gz Summary statistics for PAD: dbGAP (accession code phs001672.v2.p1) Summary statistics for carotid plaque: https://www.ncbi.nlm.nih.gov/projects/gap/cgi-bin/study.cgi?study_id=phs000930.v6.p1; accession phs000930.v6.p1 Cross-ancestry summary statistics for coronary artery calcium: http://ftp.ebi.ac.uk/pub/databases/gwas/summary_statistics/GCST90278001-GCST90279000/GCST90278455/

Individual-level scRNA-seq data from human carotid plaques was downloaded from Gene Expression Omnibus accession number <u>GSE260657</u>

## Data Availability

All data produced in the present work are contained in the manuscript

https://www.science.org/doi/10.1126/science.abf3041

https://eqtlgen.org/sc/datasets/1m-scbloodnl-eqtls.html

https://www.eqtlgen.org/cis-eqtls.html

http://ftp.ebi.ac.uk/pub/databases/gwas/summary_statistics/GCST90104001-GCST90105000/GCST90104538/

https://www.cardiogramplusc4d.org/media/cardiogramplusc4d-consortium/data-downloads/UKBB.GWAS1KG.EXOME.CAD.SOFT.META.PublicRelease.300517.txt.gz

https://www.ncbi.nlm.nih.gov/projects/gap/cgi-bin/study.cgi?study_id=phs000930.v6.p1

http://ftp.ebi.ac.uk/pub/databases/gwas/summary_statistics/GCST90278001-GCST90279000/GCST90278455/

https://www.ncbi.nlm.nih.gov/geo/query/acc.cgi?acc=GSE260657

