## Supplementary figures for "Single-cell transcriptome-wide Mendelian randomization and colocalization analyses uncover cell-specific mechanisms in atherosclerotic cardiovascular disease"

**Supplementary Figure 1.**

(a) Bar graph of the number of genes analyzed for each cell type for all outcomes in the discovery Mendelian randomization

(b) Stacked bar graph of the percentages of numbers of SNPs used in instrumental variables for discovery Mendelian randomization analysis of all gene/cell-type/outcome combinations

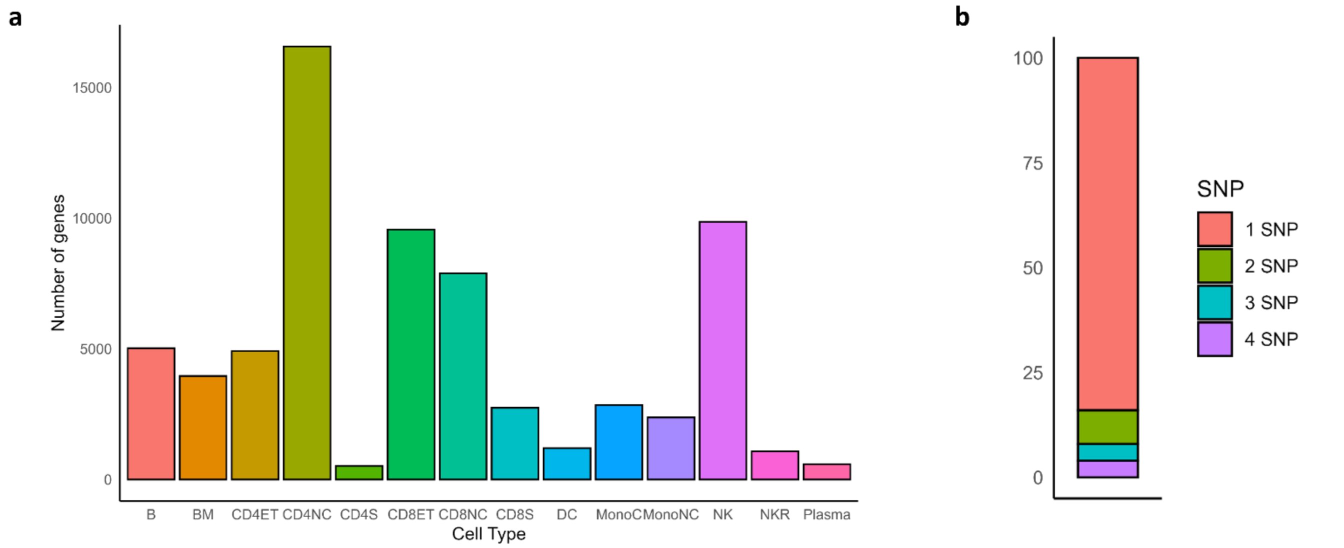

**Supplementary Figure 2** Stacked bar graph of the number of (a) eQTLs (b) genes unique to single-cell analyses

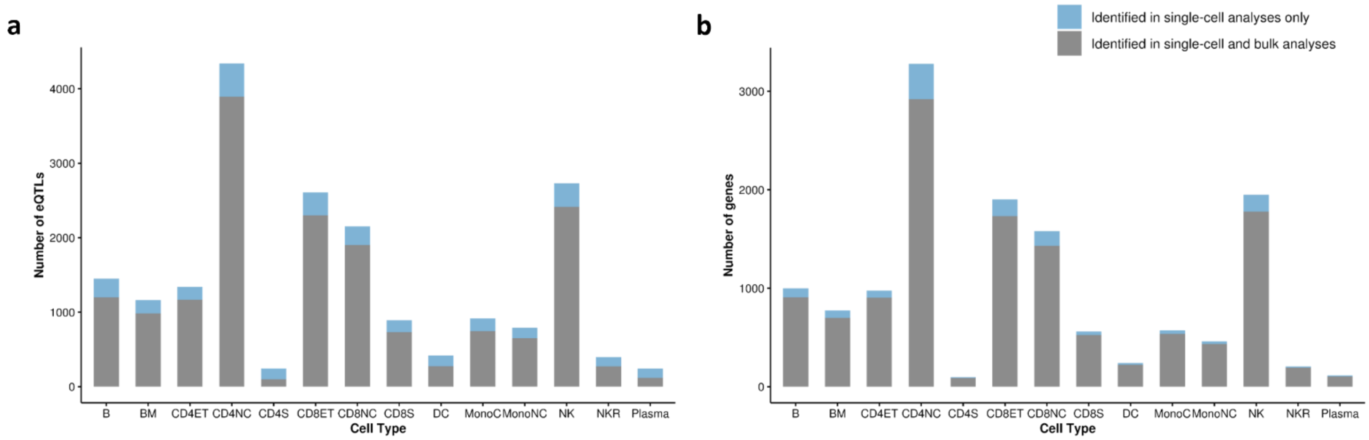

**Supplementary Figure 3.** Uniform Manifold Approximation and Projection (UMAP) of scRNA-seq data of 15 human atherosclerotic plaque samples.

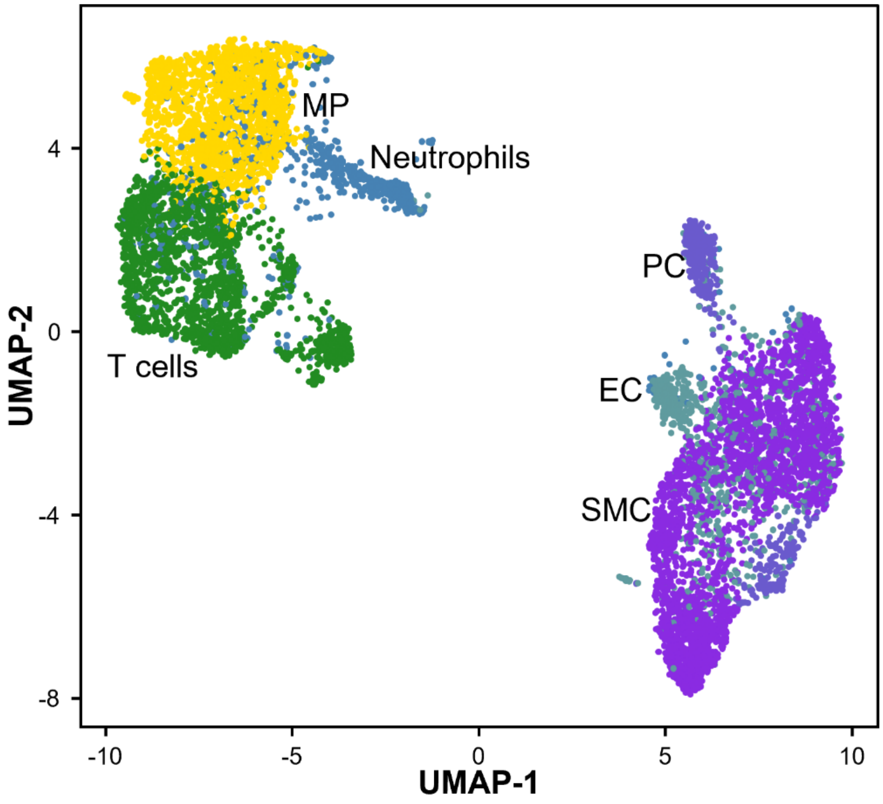

**Supplementary Figure 4.** LIPA in macrophages (CD68) in human carotid plaque sections from symptomatic patients (n =3).

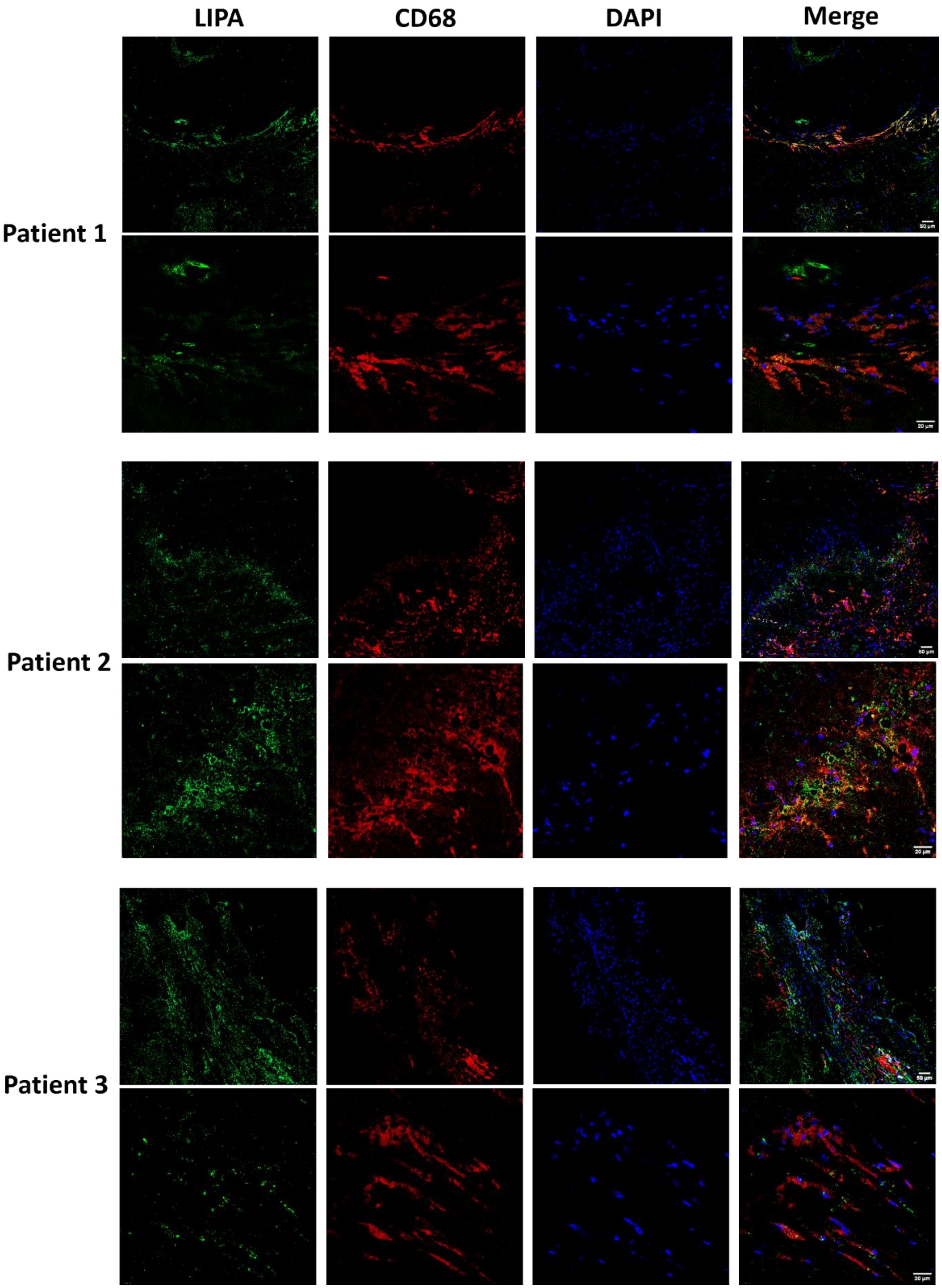
